# A Pragmatic Trial of a Multilevel Community-Centered Intervention to Reduce Pregnancy Related and Associated Morbidity and Mortality and Disparities: A Study Protocol

**DOI:** 10.64898/2026.07.23.26358783

**Authors:** Cristian I. Meghea, Hannah Bolder, Xiao Yu, Jennifer E. Johnson, Kimberlydawn Wisdom, Jaime Slaughter-Acey, Margaret Vander Meulen, Celeste Sanchez Lloyd, Jennifer Raffo, Hannah Nelson, Ran Meng, Lee Anne Roman

## Abstract

**Introduction:** Maternal morbidity and mortality in the US disproportionately affect some women, including the Medicaid-insured, rural, and Black, Hispanic, and Native American populations. The drivers of disparities are complex and there is an urgent need for multilevel interventions focused on pregnancy-related and associated morbidity and mortality (PRAMM). This study will be the first large-scale test of Community Health Worker (CHW) inclusive home visiting and provider/practice/system level improvement strategies that support empowered patients, pro-active providers, and integrates community and clinical care designed to reduce PRAMM and disparities among women who are disproportionately affected.

**Methods and analysis:** The primary study outcome will be rates of PRAMM – a composite outcome including pregnancy-associated and pregnancy-related morbidity, severe maternal morbidity, and mortality –and disparities among women disproportionately affected. A quasi-experimental, stepped wedge design will be used. Participants will be Medicaid-insured women in three Michigan counties observed during pregnancy, at birth, and up to one year postpartum, who give birth between 2021-2028 (>101,000 births). Individual health outcomes will be assessed at three steps of the stepped-wedge design. Analyses will use a statewide linked data system including all Medicaid birth and death records, Medicaid claims, and other program data. In the context of Michigan extending pregnancy-related Medicaid, the study will evaluate the impact of the proposed multilevel intervention vs. usual care from early pregnancy through 12 months postpartum. The study was developed with full community engagement and participation including shared leadership with academic and community-based Principal Investigators representing intervention communities.

**Ethics and dissemination:** An Institutional Review Board determined the study exempt from the human subject’s regulations, for the following reasons. Two of the three intervention components will not interact with human subjects to collect data - we will only rely on secondary research using data or biospecimens not collected specifically for this study, and the data will be provided without identifiable information by someone without any role in this research study except providing the data. The third intervention component, medical provider surveys, is a benign behavioral intervention where information will be recorded using methods that prevent subjects’ identities from being readily ascertained. Results will be published in peer-reviewed journals following EQUATOR guidelines.

**Strengths and limitations of this study:**

- This large pragmatic trial will address disparities that occur at multiple levels – community, provider and systems – and will leverage strong existing partnerships with community groups.
- The study’s rigorous step-wedge design will enable evaluation of effectiveness in reducing maternal morbidity and mortality rates and disparities among women disproportionately affected, as well as tests of mechanisms and cost-effectiveness.
- Limitations include the evaluation focus on the Medicaid insured population, which may reduce generalizability.

## INTRODUCTION

The US maternal mortality rate is the highest among developed countries.^1-3^ Around 80% of the deaths are . preventable^4^ Of all deaths, approximately one-third occur between one week and one year postpartum, with one-firth of deaths occurring between seven and 42 days postpartum^5^. Severe maternal morbidity (SMM), considered proximate to maternal mortality, affects about 60,000 US women every year during pregnancy, delivery, and the first six weeks postpartum.^2,6,7^The Center for Disease Control (CDC) defines SMM as “unexpected outcomes of labor and delivery that result in significant short- or long-term consequences to a woman’s health.”^8^ The US SMM rate increased 40% between 2016-2021,^9^ higher than in other high-income countries,^10^ affecting mothers, families, and communities, and costing billions of dollars per year.^11^

Non-Hispanic Black (hereafter, Black) women in the US are three to four times more likely to die of pregnancy related complications ^7^ and have twice the rates of SMM,^2,12^ with the highest rates for 22 of 25 SMM indicators used by the CDC^7^. Black women also have higher rates of other conditions, including elevated rates of perinatal depression and anxiety disorders,^13,14^ hypertension,^15^ preeclampsia,^16,17^ and delivery complications,^7,18^ conditions.

The persistence of disparities in rates of pregnancy related and associated morbidity and mortality (PRAMM) has led to calls for comprehensive, multilevel approaches at the individual, provider/practice, community, and system level^7,20,21^. While efforts have focused on the individual level, strengthening the community environment of pregnant and postpartum women with additional support, availability of services, and connection with care and community resources, could potentially impact PRAMM - especially for women with complex care needs. At the provider/practice level, improving provider communication, enhancing the content and processes of care for those at greater risk, and developing clinical and community health linkages for coordinated care may improve care.^22,23^ At the system level, connections between health care and local community organizations, who understand the unmet social needs of pregnant and postpartum women, are necessary to improve safety and quality care.^24^

There are also increasing demands for community-centered care, including Community Health Workers (CHWs), who are uniquely positioned to reach high-risk populations with medical and social needs.^25^ A CHW is “a frontline worker who is a trusted member of a community and has a thorough understanding of the community being served.”^26^ Evidence both from US^26^ and from low/middle-income countries^27^ show that CHWs can improve maternal care and health by using shared experiences and knowledge to build trusting relationships with pregnant and postpartum women. In general, CHWs focus on care access, quality, and social determinants of health (SDOH) in the following ways: empowerment and peer support, system navigation of health resources, connections to care and supporting treatment; facilitation of access to resources (food, housing, transportation), use of problem solving and coaching for client-determined goals, and specific health education curriculums to promote wellness and address risk factors.^28^

Perinatal CHW-inclusive home visiting is uniquely positioned to reach populations with pre-existing comorbidities (e.g., hypertension and diabetes), empower women to manage pregnancy complications and chronic conditions more effectively, navigate the healthcare system, and address SDOH. Recent evidence indicates CHW-inclusive home visiting, in a team approach with nurses and social workers, reaches the highest risk women who are disproportionately affected, improves prenatal and postnatal care, and reduces the risk of and disparities in adverse birth outcomes.^29,30^ Further, this CHW-inclusive home visiting program was associated with an increase in visits and penetration in segregated neighborhoods, compared with home visiting without CHW involvement.^31^

While CHWs can be an important component of care, disproportionately affected pregnant and postpartum women are more likely to be reluctant to ask questions in healthcare settings,^32^ experience barriers to open communication,^33^ decline care during the childbirth hospitalization,^34^ and are less likely to attend a postpartum check-up.^35^ Thus, at the provider and practice level, strategies are needed to address social and environmental stressors and help providers implement best practices for maternal health (e.g., life-saving patient-provider communication, provider advocacy to connect high-risk women with community-based care).

To address persistent disparities in PRAMM, the study will evaluate the effectiveness, mechanisms and cost-effectiveness of a multilevel intervention designed to reduce PRAMM disparities in disproportionately affected women, including Medicaid, Black and Hispanic populations. This study will be conducted as one of three studies in a US National Institutes of Health (NIH) Maternal Health Research Center of Excellence (The Multilevel Interventions for Maternal Health and Disparities – MIRACLE - Center) as part of NIH’s Implementing a Maternal Health and Pregnancy Outcomes Vision for Everyone (IMPROVE) initiative. The intervention will include community and provider/practice level strategies with implementation in three Michigan communities. The study was developed with community engagement and participation to privilege community voices from study inception and includes shared leadership with academic and community Principal Investigators (PIs) from each of the three geographical intervention sites. At the community-level, a CHW-inclusive home visiting model will be adapted to increase pre- and postnatal focus on maternal health, pregnancy complications, and chronic conditions. At the provider/practice-level, the study will provide actionable, experiential activities to address patient-provider communication, and awareness of community care services including CHWs and home visiting (e.g., meet SDOH needs). In addition to these two levels, at the system level, the 3 communities will be participating in another Center related study focused on the implementation and testing of Alliance for Innovation on Maternal Health – Community Care Initiative (AIM-CCI) maternal safety bundles that explicitly focuses on PRAMM disparities.

Specifically, the aims of the study are to: (1) to assess the effectiveness of the intervention (vs usual care) in reducing PRAMM (up to 1-year postpartum, overall, and disparities) and the PRAMM subset of SMM and pregnancy-associated mortality (up to 1-year postpartum, overall, and disparities); (2) to test access to care, quality, and social conditions as mechanisms of the effect of the multilevel intervention on PRAMM and disparities; and (3) to evaluate the cost-effectiveness of the multilevel intervention.

The study is innovative in that it (1) is the first large-scale test of a scalable and integrated CHW home visiting program specifically designed for reducing PRAMM and disparities among women disproportionately affected, including Medicaid, Black and Hispanic women; (2) will include interventions for specific comorbid conditions relevant to PRAMM and disparities, including multimorbidity; (3) will be one of the first to address patient-provider interactions both from the provider and patient perspective; (4) and will enhance the coordination of clinical and community care at multiple levels.

## METHODS AND ANALYSIS

### Study Setting and Population

The study will examine outcomes among Medicaid-insured populations because: (1) Medicaid covers a disproportionate share of women at risk for adverse perinatal outcomes;^36-39^ (2) Medicaid beneficiaries have greater odds of PRAMM compared to privately-insured women;^10,40^ and 60% of maternal deaths in Michigan occurred to women with Medicaid insurance;^41^ (3) Medicaid, in Michigan, is interested in the outcomes of this trial as it will inform CHW costs and policies for potential scale up in high-risk populations.; and (4) Medicaid covers nearly half of all US births (43%; 41% in Michigan)^38,42,43^ and more than half among women disproportionately affected by PRAMM (e.g. 66% of Black births). In addition, Michigan Medicaid has extended pregnancy-related coverage to 12 months postpartum beginning in 2022. This presents an extraordinary opportunity for the study intervention to impact PRAMM and disparities continuously from pre- to postpartum. Women will participate in the intervention from early pregnancy though 12 months postpartum; study data will allow us to observe the entire pregnancy, including follow-up at birth, through the postpartum year.

The intervention will be deployed and assessed for effectiveness and cost-effectiveness in Wayne County (including Detroit), Genesee County (including Flint) and Kent County (including Grand Rapids, the second largest Michigan metro area). The study chose these three counties to test the effectiveness of the proposed intervention because: (1) Wayne County is the most populous Michigan county, and includes Detroit, where women experience some of the largest PRAMM disparities in the nation; (2) there are well-established partnerships and ongoing collaborations, including the Michigan State University-Henry Ford Health (MSU-HFH) strategic partnership, in these counties that ensure feasibility of the study, especially deploying the multilevel intervention; (3) the large Henry Ford Health system and the CHW network could aid in further scale-up if the multilevel intervention is found effective; (4) the three counties represent different kinds of communities (i.e., East vs. West Michigan).

Many states have home visiting programs with universal Medicaid pregnancy-related eligibility and an existing statewide service infrastructure. In Michigan, MIHP (Maternal Infant Health Program), is the statewide evidence-based,^36,37,44^ Medicaid-sponsored home visiting program delivered by licensed professionals. Under a Michigan Pay-for-Success initiative, a CHW-inclusive home visiting team model, called Strong Beginnings, was tested with CHWs teamed with MIHP nurses and social workers to reach and serve Medicaid-insured pregnant/postpartum women. While all Medicaid-insured women who were pregnant, living in service area, and desired to enroll in services were eligible, interest was highest among women disproportionately affected by poor maternal health outcomes. Participation in Strong Beginnings was found to reduce the risk of preterm and very preterm birth with significantly larger effects among Black women and to improve both access to and quality of perinatal care (e.g. timely postpartum checkups).^30^ Building on these supportive findings, the current study will test the effectiveness of a CHW-inclusive home visiting program specifically focused on PRAMM and disparities on a much larger scale.

### Study Design

The study will run 07/01/2023 – 06/30/2030. The use of quasi-experimental stepped wedge design, as in Figure 1, will result in all three counties (Wayne, Genesee, Kent) receiving the intervention, which is key for stakeholders. To balance the steps with respect to size and to allow sufficient resources for intervention delivery, Genesee and Kent counties will form the first step and Wayne County, larger in size, will be the second step of the stepped wedge design. Randomization of the order of cross-over would be desirable but not possible here.

**Figure 1.**
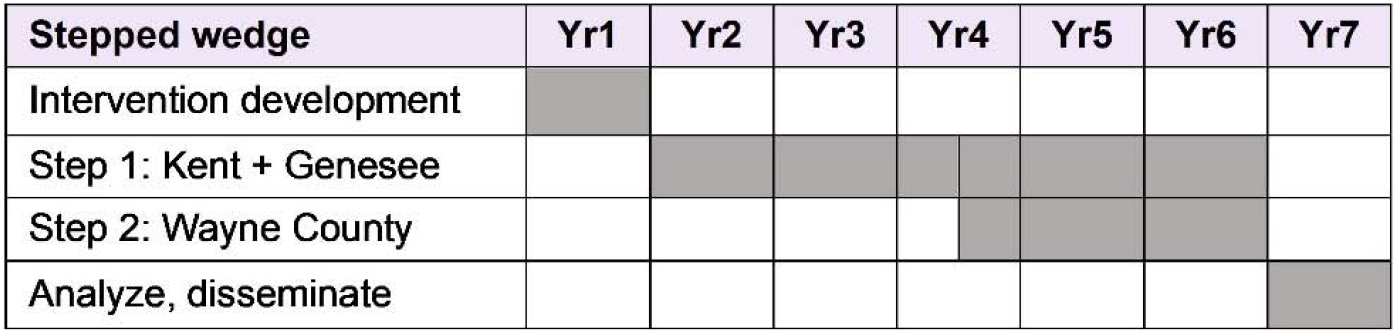
Timeline for this quasi-experimental stepped-wedge study: intervention development, intervention (steps 1 and 2), and data analysis and dissemination

In addition to traditional prenatal care, Medicaid-insured pregnant and postpartum women in Michigan are eligible for MIHP services, available in most Michigan counties. However, statewide, only about 28% of Medicaid-eligible women enroll in the program with participation of one in four Black and one in five Hispanic beneficiaries.^45^ The study will utilize a MIHP prenatal indicator to identify women in the data who participate in MIHP.

### Intervention

The conceptual model framing the intervention and addressing how levels of influence (community, provider, and system levels) and domains of influence (care access, quality, social conditions) intersect and interact, is based on Howell’s model disparities in severe maternal morbidity and mortality^7^ and aligns with the US National Institute on Minority Health and Health Disparities (NIMHD) Research Framework. Pregnant and postpartum women are impacted at multiple levels, including community, provider, and system levels. Further, improved quality and access to care and improved SDOH conditions mediate the relationship between community, provider, and system-level factors and PRAMM and disparities. Figure 2 shows the model that informs the study and intervention design.

**Figure 2.**
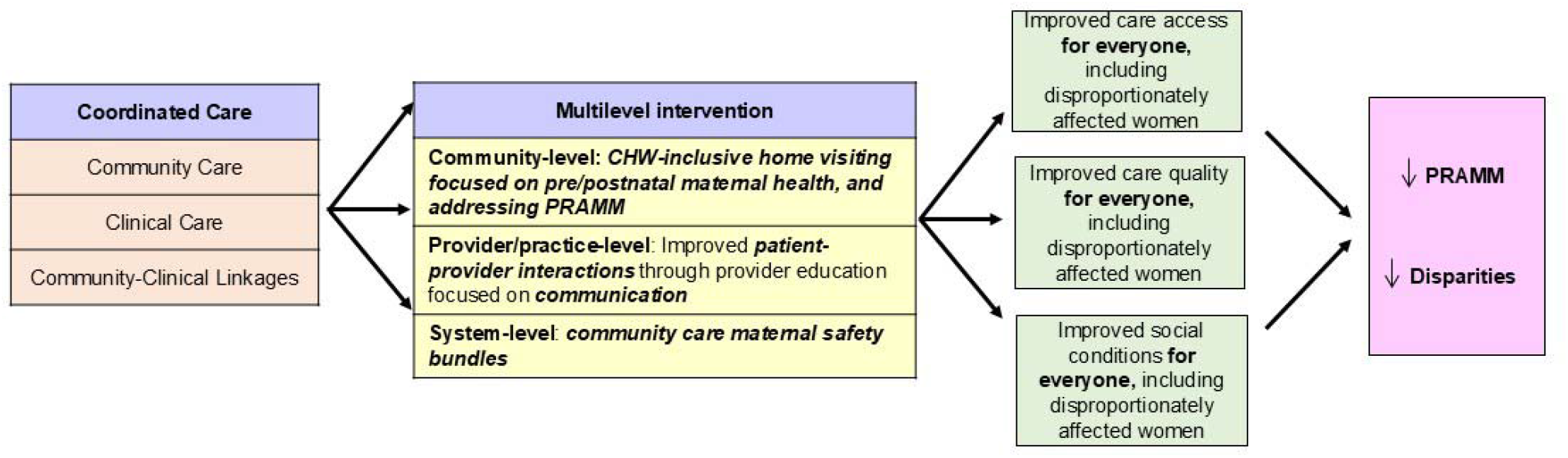
Conceptual model of how the multi-level intervention may decrease Pregnancy-Related and Associated Morbidity and Mortality (PRAMM) and disparities

This study will test a multilevel intervention. First, at the community-level, a CHW-inclusive home visiting model will be specifically adapted to increase pre- and postnatal focus on maternal health and to address PRAMM and disparities among women disproportionately affected. Second, at the provider/practice-level, actionable and experiential trainings will address provider communication (e.g. listening to pregnant and postpartum women), and linking clinical care to community care services including CHWs and home visiting (e.g. how they address SDOH), for both clinical and community settings. Third, at the system level, a MIRACLE Center companion project^46^ will scale-up prior implementation efforts in synergy with this project and will test maternal safety bundles for community care that are explicitly focused on disparities. A factorial design will enable evaluation of these complementary components.

### Community Level Intervention

The community-level CHW-inclusive home visiting model will include interventions for PRAMM-relevant comorbid conditions such as hypertension and diabetes (e.g. self-managing chronic disease and multiple conditions, adhering to medical appointments and care) from early pregnancy through the first year postpartum. The study’s community-level intervention will enhance CHW’s role on the care team by empowering women to be full participants in their care (e.g. recognize warning signs, engage with providers to have life-saving plans in place). Specifically, CHW-inclusive home visiting will include interactive CHW learning interventions designed for home visiting: (1) to understand SMM/PRAMM risks and recognize warning signs and critical life-saving actions; (2) to seek timely care and engage providers/and practices in potentially life-saving conversations; (3) to discuss with providers maternal risks and access to resources that can mitigate negative effects; (4) to have advocacy skills to engage providers/practices in improving their care; and (5) to successfully transition from prenatal care to primary care for interconception care. The community level interventions will be manualized, including the content and frequency of CHW visits, the focus on early prenatal enrollment, and health and social support interventions focused on reducing PRAMM and disparities.^29,36,37^

The added CHW interventions are designed to improve patient satisfaction, increase self-advocacy, improve early and timely service utilization, and address SDOH as they relate to PRAMM. By adapting a home visiting model to include CHWs, increase the postpartum focus on PRAMM and disparities; and empowering women as it pertains to their PRAMM and disparities, it is anticipated that there may be aligning of existing community care services to better respond to community realities. ^40^

### Provider/Practice Level Intervention

It would be burdensome and frustrating to empower Medicaid-insured women to address PRAMM without also driving changes at the provider, practice, and system levels.^23^ At the provider/practice level the study will address communication gaps through experiential, actionable, maternal health provider strategies and education. Selected strategies will be informed by the collaborators’ prior work in developing and implementing maternal mortality education, developed in partnership with women from the target communities with decades of experience in addressing maternal and infant health disparities,^47-49^ and other work with obstetrics and gynecology physician residency teachers.

Educational initiatives will include maternal morbidity and mortality causes and disparities, stressors in medical practices and communities, strategies to implement best practices linking clinical and community care and resources, and help providers better identify how to address maternal health. Skill building will include how to engage in life-saving conversations (e.g., increased risk, warning signs), empower women as partners in care (e.g., shared decision-making), and engage patients in community programs and resources. Emphasis will be on functioning in team-based approaches for integrated SDOH care and referrals, as well as advocating with patients for participation in community-based perinatal home visiting programs and other community resources. With the collaboration of providers, patients, and community partners, the study will also develop mechanisms and protocols for to address contributing factors of maternal morbidity and mortality. These protocols will address pregnancy complications and chronic conditions, treatments, follow-up processes, standardizing referrals, and timely access for home visitors to practice/providers. The latter is especially important in coordinating care for women at greater risk for PRAMM and with complex care needs.

### Systems level Intervention

Previous efforts have shown that use of hospital-focused maternal safety bundles is an important part of successful efforts to reduce PRAMM. Informed by this success, the national AIM-CCI initiative developed quality improvement bundles that focus on (1) PRAMM disparities and (2) community care (i.e., care provided outside of the hospital in outpatient and other community settings) and coordination among care settings. Kent and Genesee Counties, national pilot community sites with AIM-CCI, previously implemented selected components of bundles - details of the implementation protocol have been described elsewhere.^50^ This implementation will be expanded in a companion project as part of the MIRACLE Center,^46^ and a factorial design will enable evaluation of these complementary components.

### Participants

The study population will include all Medicaid-insured women during pregnancy, at birth, and in the 12 months postpartum, who gave birth between January 1, 2021-December 31, 2028 (∼101,000 births), as identified in the Michigan Department of Health and Human Services (MDHHS) data warehouse. This warehouse includes complete Medicaid pregnancy-birth-postpartum cohorts, including prenatal, pregnancy, and postnatal mother Medicaid claims linked to enhanced prenatal care screening data, as well as birth and death records. Maternal race and ethnicity will be assessed based on birth records. Mothers with multiple choices will be coded, in this order, Hispanic if they chose this ethnicity, and Black if not Hispanic and one of the choices was Black race.

Mothers will be coded as White if they did not choose Black race or Hispanic ethnicity and chose White race. County of residence at delivery (Wayne, Kent, and Genesee), will be used as a criterion for inclusion in the analytical sample. County of residence at delivery will be used to determine the exposure to intervention based on the condition (experimental or control) of the county at the time of pregnancy and postpartum period. The sample of women for each step of the stepped wedge design will be selected so that the time of county cross-over to experimental condition is not included in the period of pregnancy, birth, and 12 months postpartum.

### Outcomes and covariates

Intervention Effectiveness: Primary and Secondary Health Outcomes The primary study outcome is PRAMM, which includes pregnancy-associated and pregnancy-related morbidity, including SMM and non-severe maternal morbidity (NSMM) ^51-52^ and mortality. PRAMM is a count variable reflecting diagnoses/procedures/incidents from NSMM, SMM, and pregnancy-related and pregnancy-associated mortality. Each of the 3 components of this composite measure are defined below. Study outcomes will include overall PRAMM rates for Black and Hispanic as well as PRAMM disparities for these groups relative to White birthing persons. The secondary study outcome is a subset of PRAMM: SMM/mortality, a yes/no outcome that = 1 if there are SMM and/or mortality events and 0 otherwise (measured overall and relative to White.

#### Component 1: NSMM

NSMM, its framework, definition, and measurement, result from the work led by the Word Health Organization’s (WHO) Maternal Morbidity Working Group (MMWG).^51^ NSMM is defined as “any health condition attributed to and/or complicating pregnancy and childbirth that has a negative impact on the woman’s well-being and functioning” during pregnancy and postpartum.^51-53^ The vast majority of NSMM are measurable using ICD-10 diagnostic and procedure codes^53^ with standard definitions adopted by the WHO.^52,53^

#### Component 2: SMM

SMM, as defined by the CDC and the American College of Obstetricians and Gynecologists (ACOG),^54^ will be assessed using the CDC’s list of 21 SMM indicators based on ICD-10 diagnosis and procedure codes^20^ that occur antepartum, intrapartum, and up to 12 months postpartum.^55^

#### Component 3: Mortality

Pregnancy-associated death will be defined as a death during pregnancy or within 1 year of delivery or termination of pregnancy,^56^ which may be from a cause related^57^ or unrelated to pregnancy and will be considered present if a death record is linked to the pregnancy or birth record. Disparities will be defined as the Black-White and Hispanic-White percent difference in outcomes.

#### Variables to Measure Proposed Mechanisms

Healthcare access will be operationalized using two indicators drawn from birth records and Medicaid claims adjusted for number of months of Medicaid coverage: (1) the Adequacy of Prenatal Care Utilization,^58^ a combination of frequency and number of prenatal visits, defined as a binary indicator (adequate: yes/no) and the total number of prenatal and postpartum Emergency Department visits. Healthcare quality will be operationalized using two binary indicators drawn from Medicaid claims and home visiting program data: (1) home visiting enrollment in the first pregnancy trimester and (2) an early postnatal visit within 3 weeks postpartum. SDOH needs will be identified using an additive score to aggregate postnatal food availability (“In the last 12 months did you or others in household ever cut size of your meals…”), housing concerns (“…have any concerns or worries about your housing situation”), and transportation access (“…have access to routine transportation”) into a score ranging from 1-3, drawn from the MIHP program data.

#### Covariates

The following variables were identified as key prognostic factors for the PRAMM and SMM outcomes: age, education, married (y/n), and chronic disease (diabetes, hypertension). These data are available for all Michigan Medicaid births via the MDHHS data warehouse. Because of the stepped wedge design, these will be equally distributed between groups of women exposed and not exposed to intervention. To provide added control for these factors and strengthen the attribution of the effects to intervention, these covariates will be controlled for in the effectiveness analyses.

## Data collection

Following the design of many current and previous studies,^44,48,49,59^ this study will rely mainly on the linked MDHHS data warehouse maintained by MDHHS. The research team has had continuous access to and works with these data through a Master Agreement between Michigan State University and MDHHS and project specific Data Use Agreements.^44^ The linked sources of data, all routinely collected by MDHHS or publicly available (i.e. census tract data), include: (1) complete Medicaid pregnancy, and postpartum (12 months after birth) medical claims, including ICD-10 diagnostics and procedures; (2) monthly Medicaid eligibility during pregnancy and the postpartum period; (3) birth records, including data on pre-pregnancy and pregnancy risk factors (e.g. chronic disease, prior preterm birth); (4) maternal death records; (5) additional program data, including income level and participation in other programs (e.g. cash assistance); (6) neighborhood and census tract data; and (7) state MIHP prenatal and postpartum screening data which includes demographics and socioeconomic status, pregnancy history, health history, family planning, prenatal care, nutrition, cigarette smoking, alcohol, other drug use, stress, mental health support, father pregnancy involvement and support, abuse and violence, and basic needs (e.g. housing). Data from sources (1) – (6) are available for all Medicaid-insured pregnant and postpartum women. Additional data from (7) are available for MIHP-screened women (∼20% of Medicaid women).

### Missing Data

Data on pregnancy-associated (including related) mortality will be complete for women with births through 12/31/2028. All women in Michigan are anticipated to retain Medicaid insurance 12 months after birth by the time the project would start, which will ensure virtually no data-induced attrition (i.e. missing claims) for NSMM, SMM, and other claims-derived measures that include the postpartum period. We anticipate minimal bias due to the fact that some women stop being Medicaid insured after birth (e.g. change to private insurance) as this pattern should be similar across intervention and control cohorts. However, we will perform sensitivity analyses comparing results with the entire sample to results including only women who will have continuous Medicaid coverage for 12 months post-birth.

### Sample size and power

The analytical sample will include ≈101,000 Medicaid insured births over the 6 years of the stepped wedge design, with data pulled over 8 years to ensure coverage of pregnancy, delivery, and 12 months postpartum within samples at each step of the study. Over 39% of the sample will be Black and Hispanic. Health outcome analyses will be at the individual participant (pregnant woman) level, at each of the 3 steps of the stepped-wedge design (pre-intervention step followed by 2 steps with sites crossing over to the intervention) the number of participants at each step will be 16,861, of whom 6,648 will be Black and Hispanic.

#### PRAMM

Even though PRAMM includes many potential conditions, each participant will have a limited number of them. Following recent data^47^, the study will assume a distribution with 70% zeros and decreasing probabilities for higher counts (.1 for 1, .04 for 2 or 3, .03 for 4 or 5, .02 for 6, and .01 for 7, 8, 9, 10) resulting in control (pre-intervention) mean of 1.03 and standard deviation 2.11 for planning purposes. The estimated intraclass correlation coefficient (ICC) was .003 based on 2019 MDHHS Health Services Data Warehouse data. The study has also generated other pre-intervention distributions that led to similar results of power analysis. Using Hussey and Hughes^60^ approach for approximately continuous outcomes in a cross-sectional design (with a new set of 5,620 participants in each observation period), 3 steps, 3 clusters, and coefficient of variation of 0.1 estimated based on the preliminary data, the study calculated power for example effect sizes (Cohen’s d), intervention minus control mean in standard deviation units in two-tailed tests at .05 level of significance.

#### SMM and mortality

For the binary outcome of SMM and mortality, based on the literature^2^, the rate of SMM is 4.35% among Medicaid-insured Black women and 2.99% among Hispanic women, and 1.3% among White women. Based on demographic characteristics in the sample (Wayne: 39.7% Black, 6.7% Hispanic; Kent: 10.7% Black, 11.3% Hispanic; Genesee 20.3% Black, 3.9% Hispanic), the control (pre-intervention) SMM rate is 4.21% for Black and Hispanic participants in the entire sample of 3 counties. Projected post-intervention rates detectable as significantly different from control were computed using binary distribution (last two rows of Figure 3; corresponding relative risk of SMM for intervention versus control for example in the table ranges 0.74-0.76).

**Figure 3.**
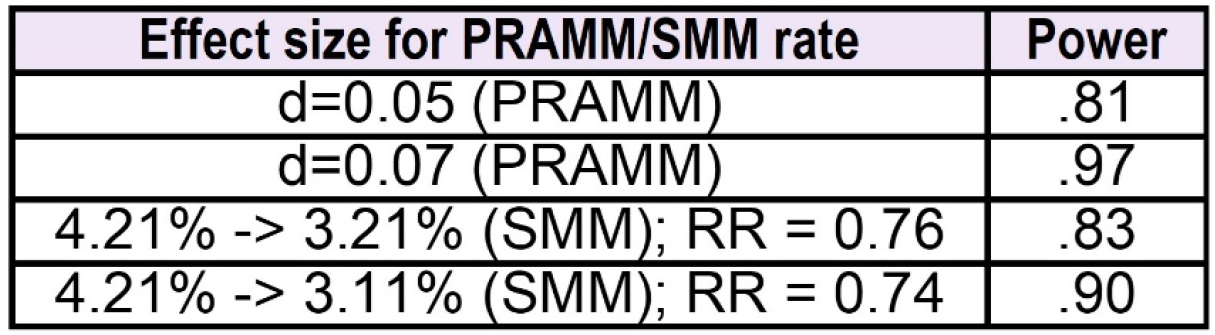
Detectable Effect Sizes for Severe Maternal Morbidity (SMM) and Pregnancy-Related and Associated Morbidity and Mortality (PRAMM) for various levels of power. Cohen’s d was used for PRAMM effect sizes and relative risk was used for SMM effect sizes.

Thus, even very small reductions in PRAMM and SMM are detectable given the proposed sample size.

### Statistical analysis

Tests will be two-sided at .05 level of significance with reported measures of clinical significance (i.e., effect sizes). The effect sizes will be estimated using differences among least square means derived by the square root of residual error from the generalized linear mixed effects models (GLMM). All analyses of effects described below will be performed for the entire sample of Black and Hispanic women or all women (to assess reduction in disparity).

Some women will no longer be Medicaid-insured after birth (e.g. change to private insurance or become uninsured). Since this pattern should be similar across intervention and control cohorts, minimal statistical bias is anticipated. However, the study will perform sensitivity analyses comparing results with the entire sample to results including only women who will have continuous Medicaid coverage for 12 months post-birth.

#### Estimation Procedures and Hypothesis Testing for PRAMM and SMM/Mortality

The PRAMM and SMM/mortality outcomes will be analyzed at the individual level to account for the unequal cluster size and to provide correct weighting for each cluster. The study will summarize and compare the demographic characteristics of participants unexposed (controls) and exposed to the intervention by virtue of delivery in a given county at a given step of the stepped wedge. By design (even in the absence of randomization), significant differences are not expected, but this will be evaluated empirically using the full data set. Correct standard errors for the parameter estimate (difference between intervention and control means) will be derived from the GLMM with appropriately distributed errors. The exponential distribution family used in GLMM allows to model skewed distributions (e.g., Gamma). GLMM will include fixed effects of time in years and intervention condition and covariates. In addition, the study will adjust for any demographic characteristics found to differ by intervention exposure group. County will be treated as a random effect to account for nesting of participants within counties; an additional random effect will be specified for any repeated pregnancies of the same woman in the data. This model will be fit first for Black and Hispanic women. Next, to test reduction in disparity by race/ethnicity, GLMM will be fit for all participants, with race/ethnicity and race/ethnicity by intervention condition interaction added. The essential parameter in this model is associated with the interaction term reflecting reduction in disparity due to intervention. The analyses of PRAMM will be followed by analyses of SMM using the same approach but with the Binomial error distribution in the GLMM. Specifically, the study will examine intervention effects for: (1) PRAMM among Black and Hispanic birthing women; (2) PRAMM disparities (Black and Hispanic vs. White); (3) SMM/mortality among Black and Hispanic women; and (4) SMM/mortality disparities (Black and Hispanic vs. White). In addition to these outcomes, the study will examine intervention effects on sub-scores of PRAMM that are based on subsets of conditions included in the PRAMM index: Black and Hispanic direct NSMM PRAMM, Black and Hispanic indirect NSMM PRAMM, and Black and Hispanic co-incidental NSMM PRAMM as well as Black and Hispanic-White disparities in these outcomes.

#### Mechanisms of Intervention Effects

To address access to care, quality of care, and SDOH as potential mechanisms for reduction in Black and Hispanic PRAMM and disparities, the study will use Preacher and Hayes approach^61,62^ to estimate direct and indirect (through the mediator) effects of intervention on PRAMM in the PROCESS macro, with county specified as a cluster. The study will consider each mediator separately (PROCESS model 2) and multiple simultaneous mediator model 4. The test of the equality of indirect effect to 0 will yield a formal test of mediation. The test will be performed using a statistical bias corrected bootstrapping analytic strategy based on 5,000 bootstrap samples to estimate confidence intervals (CIs) around the indirect effect of intervention on the outcome. To establish mediation, the 95% CI around the indirect effect must not include 0.

#### Cost-effectiveness/Cost-benefit Outcomes

The study’s grant accounting will capture the costs of providing the multilevel intervention. The primary cost-effectiveness (CE) measure will be NSMM. For NSMM, intervention costs per point of NSMM score reduction will be calculated. Secondary CE measures will be maternal mortality and SMM. Prevented SMM will also be monetized using Medicaid claims data; this will be done by calculating the difference between Medicaid delivery expenditures between women with SMM and without SMM, using the study’s claims data and prior estimates. The value of a statistical life (VSL)^63^, currently around $10 million, will be used to monetize prevented maternal deaths. Costs (and savings) in future years will be discounted to present value in the year of treatment initiation using a 5% rate (and 8% in sensitivity analysis).

### Strengths and limitations of study design

The research design leverages strong existing partnerships with community groups, and the whole study design process – from generation of research questions, intervention design, to evaluation and dissemination – was a direct product of discussions and collaborations with the affected communities. This research was not merely informed by community input but was generated by community members in collaboration and partnership with the PIs.

Tested in the context of systemwide community care maternal safety bundles, this will be the first study to test the effects of a community-centered multilevel intervention that includes (1) CHWs teamed with licensed providers embedded in a statewide program and (2) actionable, experiential, provider trainings focused on communication to improve patient-provider interactions. The evidence generated by this study has significant potential to be adopted by public health policymakers and programs (e.g., Medicaid CHW reimbursement), and implemented in clinical practice to reduce disparities in maternal mortality and morbidity.

### Data Management

The Honest Data Broker will be the only person accessing personal identifiable data in the MDHHS data system. They will generate de-identified unique individual record numbers, remove key personal identifiers, and provide the research team with limited (per HIPAAA definitions) data. The de-identified unique individual record numbers will allow the research team to link the relevant records at the individual mother and infant level. Data will be stored on a secure server, accessible only by the Michigan State University data analysis team with the use of a VPN.

### Patient and public involvement

This study is part of the NIH-funded MIRACLE Center, which has a Community Partnership Component (CPC) whose purpose is to provide feedback about Center projects and to embed stakeholder voices into the direction and operation of the Center as a whole. Community members participated in research planning, study design, and development of the intervention, including burden assessment. These community members will be active at every stage and level of the research process, including the work-planning, intervention implementation processes, data analysis, and dissemination. This study includes both academic and community PIs and follows the principles of community-engaged research.

Specifically, the development of the CHW-inclusive home visiting intervention was grounded in a long-standing partnership with the Strong Beginnings federal Healthy Start program, who with 100 other US program sites, has pioneered CHW models for over 30 years. Over time, to privilege CHW voices, CHWs have co-led focus groups, conducted surveys and interviews, coauthored publications, and were actively engaged in the intervention development and implementation. Provider interventions, grounded in earlier work developed and implemented by community stakeholders, will be enhanced with provider collaboration and leadership.

Partners in the 3 communities represent networks that can be leveraged for dissemination and diffusion. While considering message length, format, message, and topic, the study will work with these partners to package interventions and findings in ways that will appeal to their members.

## Data Availability

We cannot share routinely collected MDHHS Health Services Data Warehouse data accessed by this project because it will not be generated by the project. Access to MDHHS data requires a data use agreement and a specific request to MDHHS from each research team. Study results will be shared widely with community, practice, and policy partners using a variety of approaches, as well as through standard academic channels.

## ETHICS AND DISSEMINATION

### Ethical considerations

As discussed above, this study follows the principles of community-engaged research, including community members as equal partners in all phases from study inception to dissemination. The Michigan State University Institutional Review Board determined the study exempt from the human subject’s regulations, for the following reasons. Two of the three intervention components will not interact with human subjects to collect data - we will only rely on secondary research using data or biospecimens not collected specifically for this study, and the data will be provided without identifiable information by someone without any role in this research study except providing the data. The third intervention component, medical provider surveys, is a benign behavioral intervention where information will be recorded using methods that prevent subjects’ identities from being readily ascertained.

### Dissemination

This trial will be published in peer-reviewed journals following EQUATOR^64^ guidelines. In addition, CPC partners will identify leaders, existing coalitions (local and national), community groups, faith-based groups, and other key stakeholders to disseminate Center findings and to create forums for discussion and engagement. CPC members will work collectively to create community focused and literacy sensitive policy briefs, community report-backs, and host community forums with local systems change leaders to cocreate systems change.

## Acknowledgements

We acknowledge the following community and clinical partners who contributed to the development of the project protocol described in this manuscript: Jaye Clement, E. Hill De Loney, James William Dearing, Kent Darnell Key, Claire Margerison, Athena McKay, Steven Ondersma, Sharon Saddler, Alla Sikorskii, and Robert Sokol.

## Notes

**Funding:** Research reported in this publication was supported by the Eunice Kennedy Shriver National Institute of Child Health & Human Development of the National Institutes of Health under Award Number U54HD113291 and by the National Institute On Minority Health And Health Disparities of the National Institutes of Health under Award Number R01MD016003. The content is solely the responsibility of the authors and does not necessarily represent the official views of the National Institutes of Health.

### Competing Interest Statement

All the authors of this study received financial support from the NIH grants U54HD113291 referenced above and Michigan State University provided some matching funds. JEJ receives consulting fees from the Michigan Health and Hospital Association for her expertise in postpartum depression prevention. This protocol study builds on the foundation of work supported by the National Institute On Minority Health And Health Disparities of the National Institutes of Health award R01MD016003, which supports research activities for the following authors: CIM, JEJ, LR, XY, HB, JER, RM, CSL and MVM.

### Clinical Trial

NCT07025057

### Author Declarations

Institutional Review Board of Michigan State University gave ethical EXEMPTION for this work

